# Analytical comparison of nine SARS-CoV-2 antigen-detecting rapid diagnostic tests for emerging SARS-CoV-2 variants

**DOI:** 10.1101/2021.05.31.21258111

**Authors:** Meriem Bekliz, Kenneth Adea, Manel Essaidi-Laziosi, Jilian A Sacks, Camille Escadafal, Laurent Kaiser, Isabella Eckerle

**Affiliations:** Department of Microbiology and Molecular Medicine, University of Geneva, Geneva, Switzerland; FIND, Geneva, Switzerland; Division of Infectious Diseases, Geneva University Hospitals, 1205 Geneva, Switzerland; Laboratory of Virology, Division of Infectious Diseases and Division of Laboratory Medicine, University Hospitals of Geneva & Faculty of Medicine, University of Geneva, 1205 Geneva, Switzerland; Geneva Centre for Emerging Viral Diseases, University Hospitals Geneva, and University of Geneva, Switzerland

**Keywords:** SARS-CoV-2, COVID-19, coronavirus, rapid diagnostic test, variant of concern, variant of interest, sensitivity

## Abstract

Several SARS-CoV-2 variants of concern/interest (VOC/VOI) emerged recently, with VOCs outcompeting earlier lineages on a global scale. To date, few data on routine diagnostic performance for VOC/VOIs are available. Here, we investigate the analytical performance of nine commercially available antigen-detecting rapid diagnostic tests (Ag-RDTs) for VOC B.1.1.7, B.1.351, P.1 and VOI P.2 with cultured SARS-CoV-2. Comparable or higher sensitivity was observed for VOC/VOI compared to a non-VOC/VOI early-pandemic virus for all Ag-RDTs.

## The study

Severe Acute Respiratory Syndrome Coronavirus 2 (SARS-CoV-2) antigen-detecting rapid diagnostic tests (Ag-RDTs) provide laboratory-independent results at the point of care and thus are powerful tools for public health interventions. Recently, clinical and analytical studies showed SARS-CoV-2 Ag-RDT detection thresholds correlate with the presence of infectious virus in symptomatic SARS-CoV-2 infections.^1,2^ However, the majority of Ag-RDT validation studies were done before SARS-CoV-2 variants of concern/interest (VOC/VOI) emerged, with the VOCs currently outcompeting earlier lineages.^3^ To date, data on routine diagnostic performance for VOC/VOIs is sparse.^4,5^ Furthermore, clinical validation studies comparing multiple VOCs in parallel are hardly feasible.

We investigated the analytical sensitivity of nine commercially available Ag-RDTs using cultured SARS-CoV-2, comparing lineage B.1.610 (first pandemic wave) with VOCs B.1.1.7, B.1.351, P.1 and VOI P.2.

Briefly, infectious titers and RNA copies of virus stocks grown in VeroE6 were quantified by plaque titration and RT-PCR (E gene), respectively. Isolates were tested in serial dilutions, starting with 5.44 Log_10_ PFU/mL, except for P.1 which had a maximum titer of 4.24 Log_10_ PFU/mL. An infectious titer of 5.44 Log_10_ PFU/mL corresponded to 10.26, 12.11, 9.86 and 11.23 Log_10_ RNA copies/mL for B.1.610, B.1.1.7, B.1.351 and P.2, respectively. For P.1, the infectious titer of 4.24 Log_10_ PFU/mL corresponded to 11.81 Log_10_ RNA copies/mL.

Ag-RDT assays were performed according to the manufacturers’ instructions, with the exception that 5 µL of virus dilution was directly added to the proprietary buffer, and then applied to the Ag-RDT in duplicates under BSL3 conditions. Results were read independently by two individuals. Any visible test band in the presence of a visible control band was considered as positive. Ag-RDT buffer without virus was used as negative control.

When analyzing results normalized to PFU/mL, comparable or better performance to the early-pandemic lineage was observed for B.1.1.7, B.1.351, P.1 and P.2 for all assays (**Figure**). Overall sensitivity and specificity for individual isolates varied between Ag-RDTs, with the best-performing assay positive at dilutions as low as 2.43 Log_10_ PFU/mL and the lower-sensitive assays positive at 4.54 Log_10_ PFU/mL. Consistently, the highest sensitivity was seen for P.1 and P.2. Although testing for analytical sensitivity with cultured virus cannot fully replace clinical data, our data provide reassuring results for the use of Ag-RDTs to diagnose VOCs. Phenotypic properties, such as a remarkable difference in the RNA:infectious virus ratio, could hint at production of defective viral particles. Their impact on diagnostic test performance should be further investigated.

**Figure.**
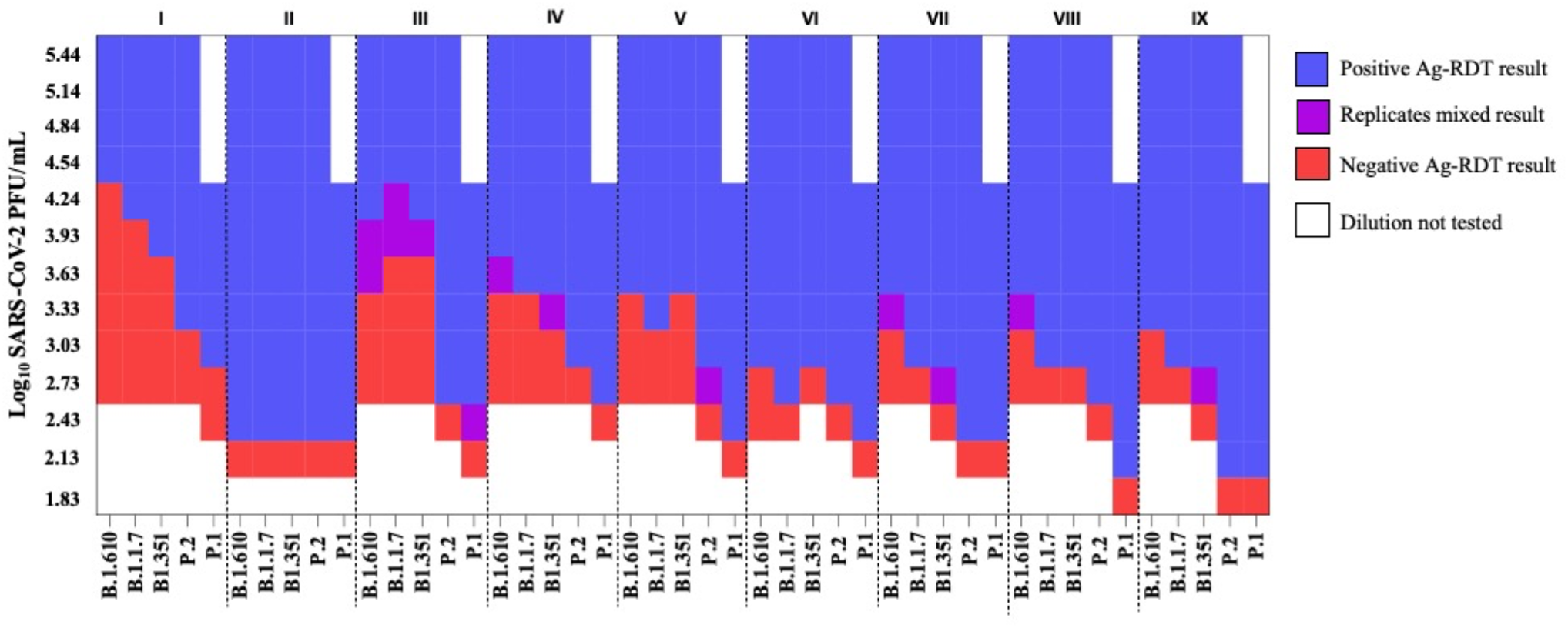
Heat map of analytical sensitivity of nine Ag-RDTs assays with SARS-CoV-2 variants of concern B.1.1.7., B.1.351, P.1 and variant of interest P.2 in comparison to an early-pandemic SARS-CoV-2 isolate (B.1.610), based on Log_10_ PFU/mL. Ag-RDTs used were I) Genedia COVID-19 Ag (Green Cross Medical Science Corp); II) Sure Status (Premier Medical Corporation); III) Joysbio SARS-CoV-2 Antigen Rapid Test Kit (Joysbio); IV) Edinburgh Genetics (Edinburgh); V) 2019-nCoV Antigen test (Wondfo), VI); Standard Q COVID-19 Ag (SD Biosensor/Roche); VII) Panbio COVID-19 Ag Rapid test device (Abbott); VIII) NowCheck Covid-19 Ag test (Bionote) and IX) Ichroma Covid-19 Ag (Boditech). All Ag-RDTs are based on detection of the nucleocapsid protein of SARS-CoV-2.

## Data Availability

Original data are available upon request.

## Acknowledgments

We thank Erik Boehm for language editing.

